# Factors associated with therapeutic complexity at the end of life: Analysis of a cohort of patients undergoing palliative care at home

**DOI:** 10.1101/2025.11.10.25339944

**Authors:** Mireia Massot Mesquida, Xavier Busquet-Duran, Josep Ma Manresa, Maria Feijoo Cid, Ramon Miralles Basseda, Pere Torán-Monserrat

## Abstract

Complexity at the end of life is the result of the interrelationship of all the patient’s needs at this stage. The Hexagon of Complexity (HexCom) enables a multi-dimensional approach to patients. The management of medication becomes a complex element for the patient/family member or caregiver.Determine the factors associated with therapeutic complexity according to the dimensions of needs determined with HexCom. Longitudinal observational study of a multicentre cohort. Therapeutic complexity was chosen as dependent variable. Independent variables: socio-demographic, relationship with the caregiver, pathologies and groups of advanced diseases, functional and mental status, six HexCom domains and diagnosis that most contributes to the death process. All these variables were assessed initially and the HexCom variables were also assessed after the patient’s death. A multivariate Poisson regression analysis was performed. The scope of the study is palliative home care in Catalonia. The cohort included 1,677 patients. Recruitment was performed by consecutive sampling. The results show a strong relationship of the initial therapeutic complexity with the difficulty in managing symptoms (OR 4.56), dying process (OR 2.62), and difficulties in handling information about the diagnosis/prognosis (OR 2.04), bad family relationships (OR 1.82) and difficulties in handling the patient’s basic needs (OR 1.87). Regarding the final therapeutic complexity, two new factors have been identified, which are the difficulty in adapting the therapeutic effort (OR 1.58) and the lack of external support (OR 0.60). This last factor is the only one which decreases therapeutic complexity as the factor increases. In order to effectively manage medication, support is needed in social, family and ethical areas, in order to reduce therapeutic complexity.

## Introduction

Polymedication is an increasingly common problem in older people with advanced chronic diseases, either adverse effects or the interactions that can occur between the different medications. Although the literature warns about the relevance of therapeutic complexity in poly-medicated patients, professionals rarely take it into account in day-to-day health care when making decisions [1–2].

From a pharmacological point of view, therapeutic complexity [3] is defined as the set of instructions, guidelines, doses and different pharmaceutical forms that a patient must take in their day-to-day life. In this regard, it can be understood that when there is more diversity in these parameters, there can be more therapeutic complexity. With the increase in life expectancy [3–4], poly-medication and therapeutic complexity are increasingly common in the population over 65 years of age.

When patients reach the end of their lives, therapeutic complexity can increase, since medicines used to ensure comfort and control of symptoms are added to the usual treatments for basic pathologies [5]. In these cases, different routes of drug administration are often combined (oral, subcutaneous, transdermal, sublingual, intranasal, etc.) which also adds complexity [5].

On the other hand, it is defined as therapeutic burden, the effort a patient must make to properly manage the medication and to take care of themselves [1,2,6,7]. It seems that the higher the therapeutic burden, the lower the medication adherence and this could be related to worse symptom control and an increased risk of hospitalizations [8–9], adding complexity to therapeutic management.

However, in patients with advanced disease and/or at the end of their lives, we should not only take into account the medication per se when we are assessing this therapeutic complexity, but will have to add to it the very nature of the surrounding circumstances which means that we are facing a paradigmatic situation of complexity [10].

The study of complexity in the care of patients at the end of their lives is currently a topic of interest, as shown in the review by Ohinata [11]. According to the systematic review by Grant et al. [12], of the 6 existing complexity classification systems, HexCom and IDC-Pal offer the most extensive determinations of complexity. The HexCom model [10] makes it possible to carry out a multi-dimensional approach towards patients at the end of their lives from all the perspectives or dimensions that influence this complexity [13–14]. This model defines complexity as “the difference that exists between the needs of the patient and the healthcare system” or that would be to say, the variance between the patient’s needs and the ability to respond to them using the available services. Therefore, it does not focus on the symptom or the suffering it generates, but on the difficulty of handling it by professionals (Fig 1)[15–17]. This complexity assessment model includes six areas of need (clinical, psychological, spiritual, social/family, ethical, and death-related needs) and is subdivided into 18 sub-areas (Table 1). It grades the complexity into three levels: low, medium or high: high complexity is considered to be when the care team cannot solve the detected need and is forced to refer the case and/or to assume that it will be limited when supporting it. This model has proven, in our environment, to be suitable for the management and inter-level referral of complex cases, regardless of the clinical type and the prognostic situation of the patient, and has demonstrated apparent validity, reliability and feasibility [18]. The present study wants to focus on the complexity related to the difficulty in the administration of the different pharmacological treatments as well as access to different techniques (Therapeutic Complexity) in palliative care with the aim of studying what factors are associated with this complexity according to the different dimensions of needs determined in the HexCom instrument (Hexagon of Complexity) in specialized palliative home care. Determining the associated factors will help us define an intervention to improve adherence that is more accurate and focused on the patient.

**Fig 1.**
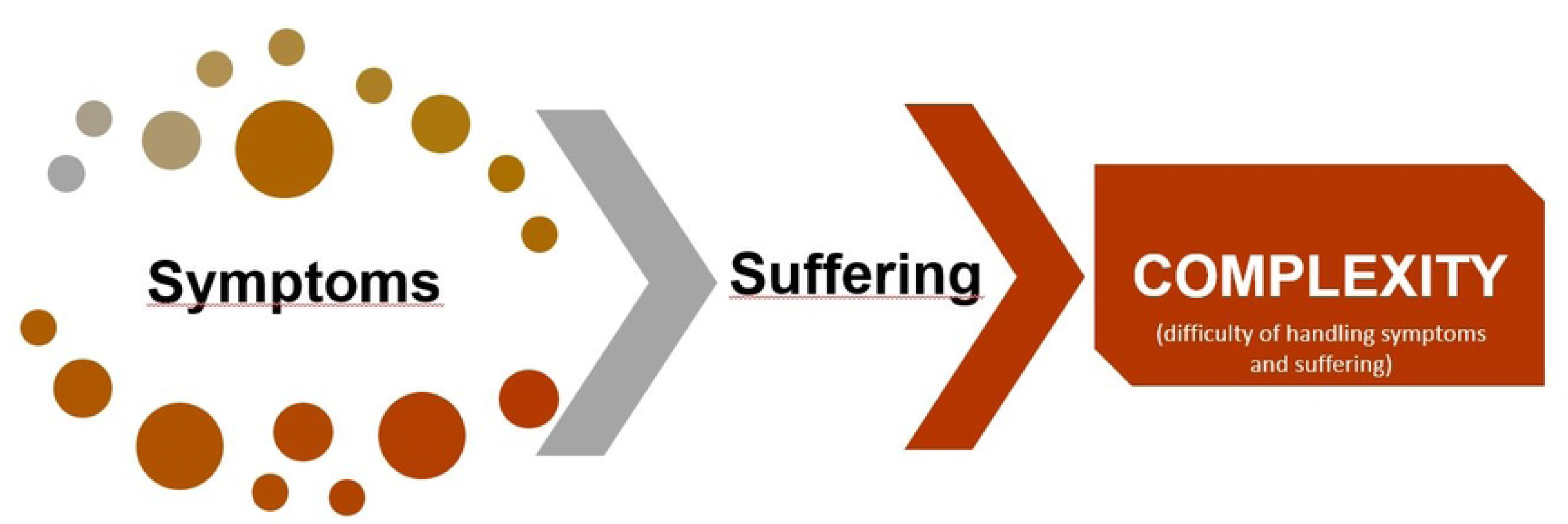
Conceptualization of complexity in the HexCom model

**Table 1.**
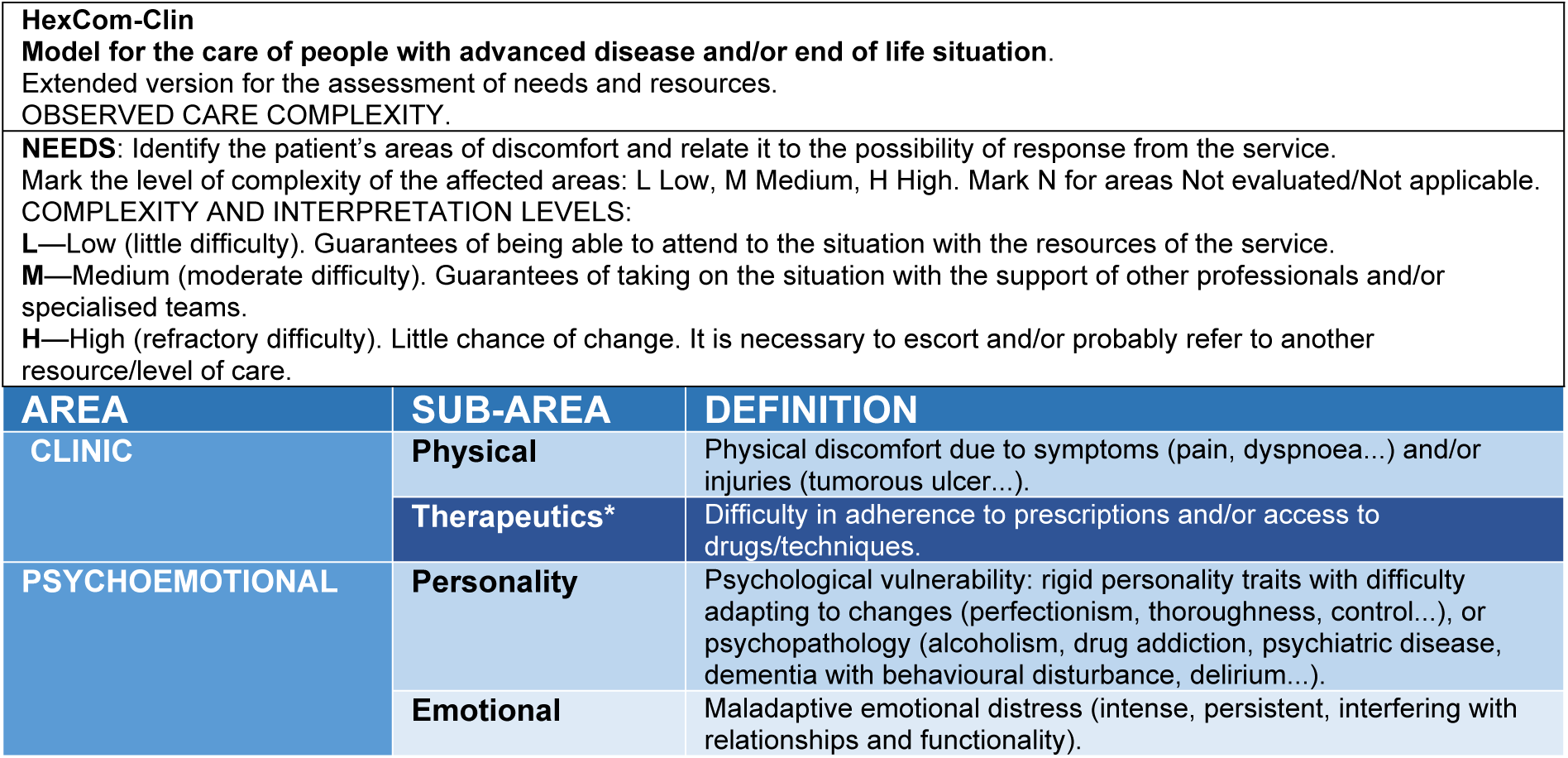
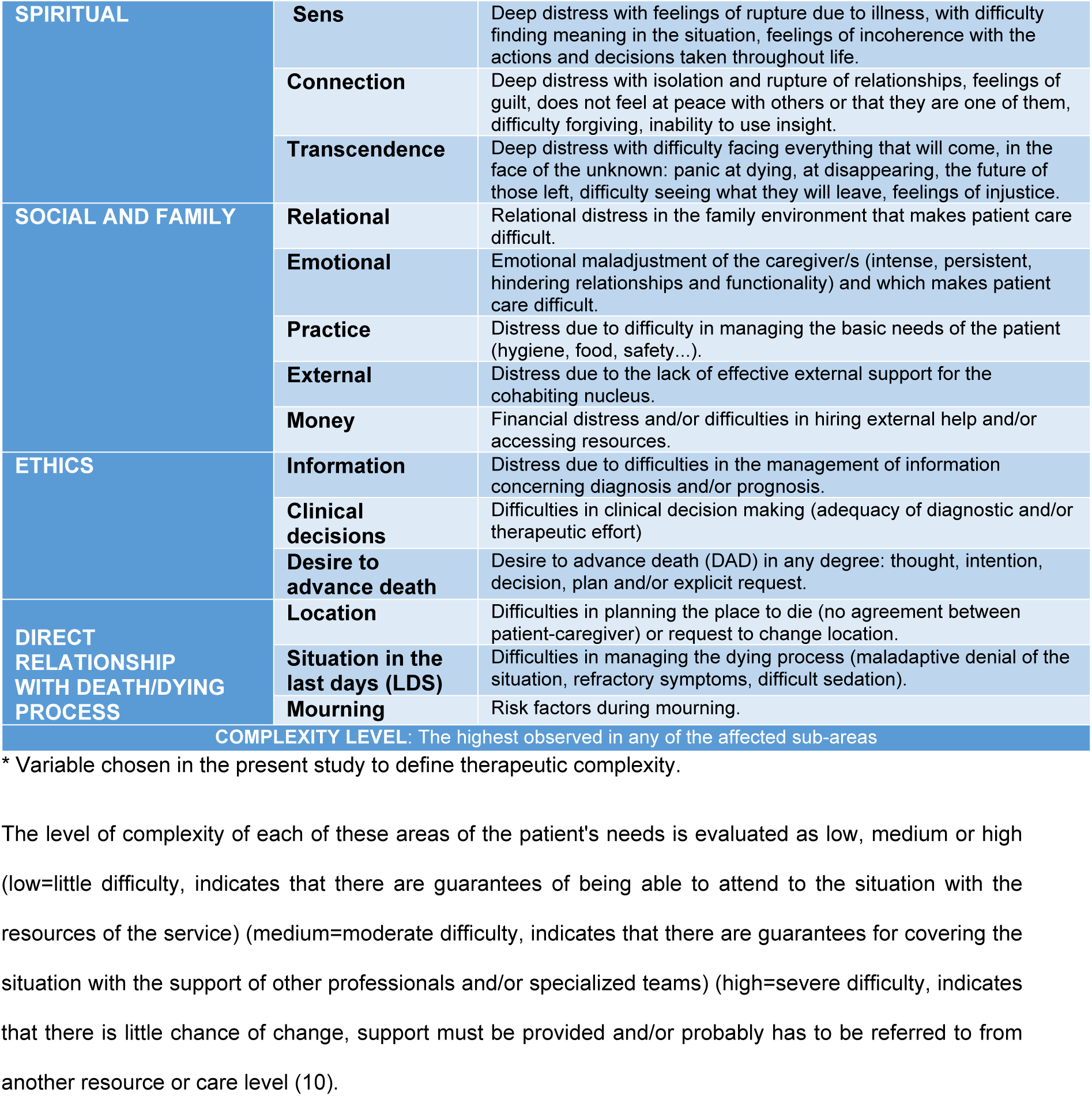
HexCom model. Six main areas of need that are assessed with the HexCom instrument [10] (clinical, psychological, spiritual, social/family, ethical and death-related needs) and the 18 sub-areas into which each of them is divided (total 18 sub-areas).

| HexCom-Clin<br>Model for the care of people with advanced disease and/or end of life situation.<br>Extended version for the assessment of needs and resources.<br>OBSERVED CARE COMPLEXITY. |  |  |
| --- | --- | --- |
| **NEEDS:** Identify the patient's areas of discomfort and relate it to the possibility of response from the service.<br>Mark the level of complexity of the affected areas: L Low, M Medium, H High. Mark N for areas Not evaluated/Not applicable.<br>**COMPLEXITY AND INTERPRETATION LEVELS:**<br>**L**—Low (little difficulty). Guarantees of being able to attend to the situation with the resources of the service.<br>**M**—Medium (moderate difficulty). Guarantees of taking on the situation with the support of other professionals and/or specialised teams.<br>**H**—High (refractory difficulty). Little chance of change. It is necessary to escort and/or probably refer to another resource/level of care. |  |  |
| AREA | SUB-AREA | DEFINITION |
| CLINIC | Physical | Physical discomfort due to symptoms (pain, dyspnoea...) and/or injuries (tumorous ulcer...). |
|  | Therapeutics* | Difficulty in adherence to prescriptions and/or access to drugs/techniques. |
| PSYCHOEMOTIONAL | Personality | Psychological vulnerability: rigid personality traits with difficulty adapting to changes (perfectionism, thoroughness, control...), or psychopathology (alcoholism, drug addiction, psychiatric disease, dementia with behavioural disturbance, delirium...). |
|  | Emotional | Maladaptive emotional distress (intense, persistent, interfering with relationships and functionality). |
| **SPIRITUAL** | **Sens** | Deep distress with feelings of rupture due to illness, with difficulty finding meaning in the situation, feelings of incoherence with the actions and decisions taken throughout life. |
|  | **Connection** | Deep distress with isolation and rupture of relationships, feelings of guilt, does not feel at peace with others or that they are one of them, difficulty forgiving, inability to use insight. |
|  | **Transcendence** | Deep distress with difficulty facing everything that will come, in the face of the unknown: panic at dying, at disappearing, the future of those left, difficulty seeing what they will leave, feelings of injustice. |
| **SOCIAL AND FAMILY** | **Relational** | Relational distress in the family environment that makes patient care difficult. |
|  | **Emotional** | Emotional maladjustment of the caregiver/s (intense, persistent, hindering relationships and functionality) and which makes patient care difficult. |
|  | **Practice** | Distress due to difficulty in managing the basic needs of the patient (hygiene, food, safety...). |
|  | **External** | Distress due to the lack of effective external support for the cohabiting nucleus. |
|  | **Money** | Financial distress and/or difficulties in hiring external help and/or accessing resources. |
| **ETHICS** | **Information** | Distress due to difficulties in the management of information concerning diagnosis and/or prognosis. |
|  | **Clinical decisions** | Difficulties in clinical decision making (adequacy of diagnostic and/or therapeutic effort) |
|  | **Desire to advance death** | Desire to advance death (DAD) in any degree: thought, intention, decision, plan and/or explicit request. |
| **DIRECT RELATIONSHIP WITH DEATH/DYING PROCESS** | **Location** | Difficulties in planning the place to die (no agreement between patient-caregiver) or request to change location. |
|  | **Situation in the last days (LDS)** | Difficulties in managing the dying process (maladaptive denial of the situation, refractory symptoms, difficult sedation). |
|  | **Mourning** | Risk factors during mourning. |
| **COMPLEXITY LEVEL: The highest observed in any of the affected sub-areas** |  |  |

\* Variable chosen in the present study to define therapeutic complexity.

## Materials and Methods

Longitudinal multi-centre observational study. The scope of the study is the community population who are in their homes and suffer from an advanced disease at the end of their lives and are cared for by specialized palliative home care in Catalonia (North-East Spain).

No randomization of patients was performed, and there was no control group. Patients were recruited in a consecutive sampling. The cohort was formed over 3.5 years, from April 15th, 2016, to December 15th, 2019. The inclusion criteria were the presence of an advanced disease in the final stage of life and the fact of being treated by the home care support teams in Catalonia (*PADES*) [19]. Patients receiving care in a nursing home were excluded.

*PADES* are multidisciplinary teams specialized in end-of-life care, made up mainly of doctors, nurses and social workers, often with the support of psychologists, who offer advice and support to primary care professionals in home care for patients with advanced chronic diseases and palliative needs. In addition to providing direct care, aimed at the control of symptoms, comfort and well-being of the sick person and their caring environment, they play an important role in the management of complex cases and in the coordination of resources between care levels [20]. A total of 45 of the 59 *PADES* teams that existed at the time (76%) participated in the study [19].

No sample size calculation was necessary given that all patients in the cohort were analysed.

The level of therapeutic complexity (high, moderate, low and not assessable) was considered as the main variable of the study (dependent variable). The patient’s socio-demographic data (age, gender, relationship with the caregiver: whether it was their partner or not), pathologies (medical diagnosis (CIE-10) and advanced disease groups: cancer, advanced chronic organ failure, advanced neurological disease, dementia or geriatric frailty/multimorbidity), patient functional status (Barthel Index) and mental status (Pfeiffer Test) were considered to be independent variables. According to the HexCom complexity hexagon: each of the six domains of needs (clinical, psychological, spiritual, social/family, ethical and death-related needs) and their 18 sub-areas were included as independent variables (see Table 1) as well as the diagnosis that most contributes to the death process. This diagnosis was determined by consensus of the *PADES* team.

To assess the therapeutic complexity, the criterion included in the HexCom instrument itself was used [10], which is defined as “difficulty in adhering to prescriptions and/or accessing drugs or techniques” (Table 1). This assessment is carried out by consensus among all the professionals within each *PADES* team.

The variable was categorized into high, moderate and low. “High” therapeutic complexity was defined as: blockage in treatment adherence. Therapeutic situation that, due to its particularities in the handling of drugs or instrument technique, may require hospital treatment or admission. Examples: intravenous drugs; paracentesis risk (septal ascites, patient with dementia, dehydration); thoracentesis; etc. “Moderate”: Difficulty in adherence to treatment. Therapeutic situation in which the handling of drugs or less common instrument techniques, or those not systematized in community health, are required. Examples: special conditions in the handling of opiates (rotation, new opiates, risk of toxicity), handling of drugs in children and adolescents or in patients with toxic habits, catheterizations, evacuating paracentesis, and “Low”: Adherence to treatment acceptable, or with guarantees of being able to undertake it despite the probable difficulties. Therapeutic situation in which drugs or instrument techniques which are common or have systematized management in community health are required. Examples: constipation due to opiates, bladder catheterization, enteral nutrition techniques, ostomies.

All variables were assessed at the beginning of the intervention by *PADES team*, through an interview with the patient and review of the clinical history (initial assessment) and also after the patient’s death (final assessment). To standardize data collection and compilation, participating *PADES* teams received 10 hours of face-to-face training, a user guide, and were given a phone number to call in case they had questions during work.

### Statistical analyses

Qualitative variables were summarized with their absolute and relative frequencies. Pearson’s Chi Square test and McNemar’s test were used in the case of analysing paired data. Given the sample size, small differences could be statistically significant, so only differences greater than 5% were considered clinically relevant.

Based on the cohort of patients included, the database was reviewed in order to proceed with its purification and analysis of the quality of the record, and a multivariate Poisson regression analysis was performed to study which factors were associated with therapeutic complexity. For this reason, the dependent variable was divided into “high/medium therapeutic complexity” and “low therapeutic complexity”. A saturated model was initially considered with all variables showing a significant association in a bivariate analysis. Next, those that were not significant were eliminated step by step (backward stepwise regression).

The significance level was 5% for all contrasts. The analyses were performed with the statistical package SPSS, version 25.0 for Windows.

### Ethical approval and consent

The study was approved by the Clinical Research Ethics Committee of the Jordi Gol University Institute of Primary Care Research (IDIAP) (registration number P15/171) and by the clinical research ethics committees of all the participating centres. All participants read and signed a written informed consent form during the PADES visit in the patient’s home.

The authors of this paper did not have access to the patients’ identifiable information. All data included in the database were pseudonimized; only the principal investigator had access to the identification code. The research team accessed the database for this study on March 1st, 2023.

## Results

A total of 1,677 patients from 45 *PADES* teams in Catalonia were included in the cohort.

Table 2 shows the characteristics of the cohort and their relationship with the existence of therapeutic complexity at the beginning (Column A) and end (Column D) of the study. Of the 1,677 initial participants (Column A) 1,555 had complete follow-up (Column C).

**Table 2.**
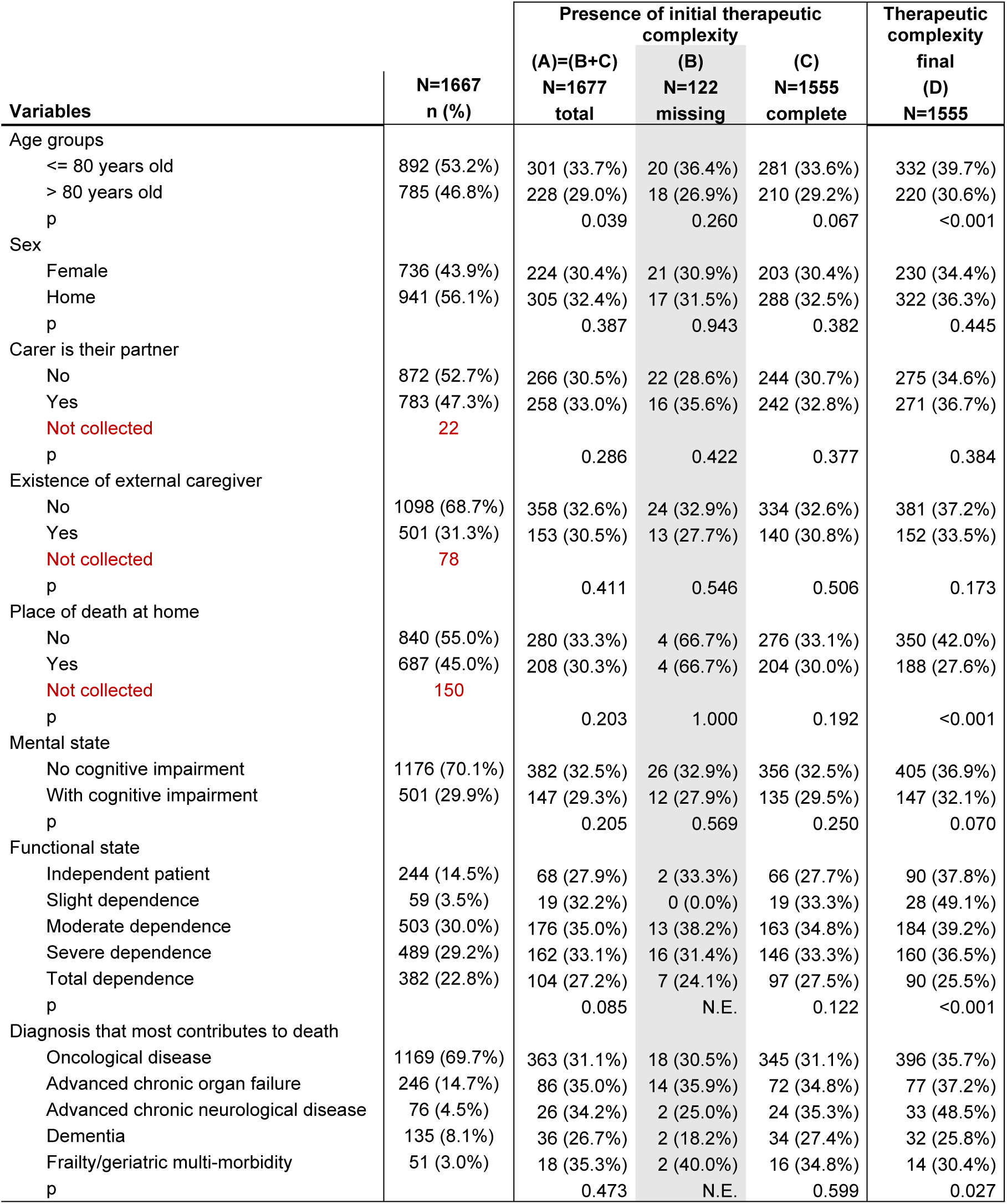
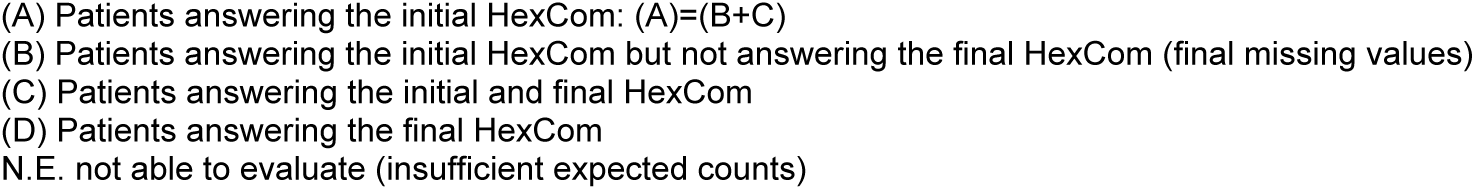
Descriptive table of the characteristics of the cohort and their relationship with the initial/final therapeutic complexity and missing values.

Of all the patients included, the initial assessment was carried out in 100% of them (Column A), while in the final assessment there was a loss of 122 patients who could not be evaluated (Column B). This loss represents 7.27% of the initial sample. In Table 2 we can see that the lost participants do not differ in any of the analysed variables. However, if we compare the complexity in the missing participants (B) compared to the complete participants (C), there are some differences greater than 5% in some variables, such as that in the missing participants a better functional state was observed and less presence of neurological disease, dementia and frailty. However, the low prevalence of therapeutic complexity in these variables presented very low expected effects and cannot be assessed statistically.

### Relationship between Therapeutic Complexity and patient variables

Initial therapeutic complexity was greater in younger patients (<80 years old) (33.7% versus 29%; p=0.039) and no other association was observed with the rest of the variables studied.

The final therapeutic complexity was also higher among those under 80 years of age (39.7% versus 30.6%; p<0.001), among those who die outside their home (42% versus 27.6%; p<0.001), among those who had a better functional capacity (independent (37.8%) or mild dependence (49.1%)) and among patients with a diagnosis of advanced chronic neurological disease (non-dementia) (48.5%).

### Relationship between Therapeutic Complexity and HexCom variables

Table 3 compares the initial and final therapeutic complexity of the included patients based on the 6 dimensions and 18 sub-areas of HexCom. The time elapsed between the initial and final assessment, considered the time of support by the *PADES* team, was 41 (14-105) days in women and 35 (14-87) days in men (p=0.07).

**Table 3.** Descriptive table of the initial and final therapeutic complexities of the cohort of patients included in the study based on the 6 dimensions of needs and the 18 sub-areas of HexCom.

| Variable* | Moderate/high complexity |  |  |  | (B*C)<br>p | (C*D)<br>p |
| --- | --- | --- | --- | --- | --- | --- |
|  | Initial |  |  | Final |  |  |
|  | total initial<br>(A)=(B+C) | Missing (B) | complete (C) | final (D) |  |  |
|  | n=1677 | n=122 | n=1555 | n=1555 |  |  |
| Clinical area | 942 (56.2%) | 71 (58.2%) | 871 (56.0%) | 930 (59.8%) | 0.640 | 0.006 |
| Physical | 833 (49.7%) | 63 (51.6%) | 770 (49.5%) | 849 (54.6%) | 0.652 | 0.000 |
| Therapeutic | 529 (31.5%) | 38 (31.1%) | 491 (31.6%) | 552 (35.5%) | 0.922 | 0.000 |
| Psychic Area | 809 (48.4%) | 44 (36.4%) | 765 (49.4%) | 728 (47.6%) | 0.006 | 0.122 |
| Emotional | 663 (39.5%) | 35 (28.7%) | 628 (40.4%) | 591 (38.0%) | 0.011 | 0.037 |
| Personality | 419 (25.0%) | 21 (17.2%) | 398 (25.6%) | 384 (24.7%) | 0.039 | 0.170 |
| Spiritual area | 519 (31.0%) | 19 (15.6%) | 500 (32.2%) | 480 (31.4%) | 0.000 | 0.344 |
| Transcendence | 311 (18.6%) | 15 (12.3%) | 296 (19.0%) | 316 (20.3%) | 0.065 | 0.175 |
| Connection | 162 (9.7%) | 8 (6.6%) | 154 (9.9%) | 162 (10.4%) | 0.228 | 0.422 |
| Meaning | 381 (22.7%) | 15 (12.3%) | 366 (23.6%) | 363 (23.4%) | 0.004 | 0.876 |
| Social and Family area | 1222 (72.9%) | 76 (62.3%) | 1146 (73.7%) | 1121 (72.1%) | 0.006 | 0.116 |
| Relational | 365 (21.8%) | 35 (28.7%) | 330 (21.2%) | 340 (21.9%) | 0.054 | 0.426 |
| Social and Family emotional | 745 (44.4%) | 51 (41.8%) | 694 (44.6%) | 731 (47.0%) | 0.545 | 0.035 |
| Practice | 634 (37.8%) | 57 (46.7%) | 577 (37.1%) | 656 (42.2%) | 0.035 | 0.000 |
| External | 883 (52.7%) | 51 (42.5%) | 832 (53.5%) | 799 (51.4%) | 0.020 | 0.014 |
| Economic | 247 (14.8%) | 19 (16.0%) | 228 (14.7%) | 236 (15.2%) | 0.699 | 0.268 |
| Ethics area | 468 (27.9%) | 21 (17.2%) | 447 (28.7%) | 390 (25.5%) | 0.006 | 0.002 |
| Desire to die | 112 (6.7%) | 3 (2.5%) | 109 (7.0%) | 112 (7.2%) | 0.053 | 0.804 |
| Clinical decisions | 295 (17.6%) | 12 (9.8%) | 283 (18.2%) | 269 (17.3%) | 0.019 | 0.395 |
| Information | 216 (12.9%) | 12 (9.8%) | 204 (13.1%) | 152 (9.8%) | 0.296 | 0.000 |
| Death area | 1067 (63.7%) | 40 (33.1%) | 1027 (66.0%) | 1064 (68.4%) | 0.000 | 0.055 |
| Location | 883 (52.7%) | 25 (20.8%) | 858 (55.2%) | 852 (54.8%) | 0.000 | 0.805 |
| Circumstances of the last days | 121 (7.2%) | 5 (4.2%) | 116 (7.5%) | 256 (16.5%) | 0.186 | 0.000 |
| Bereavement | 543 (32.4%) | 20 (16.7%) | 523 (33.6%) | 556 (35.8%) | 0.000 | 0.042 |

\* Description of variables Table 1

In the initial assessment 491 (31.6%) patients showed a high-moderate therapeutic complexity, while in the final assessment there were 552 (35.5%: p<0.001) (Column B*D), showing a slight increase in therapeutic complexity during the process.

If we compare the initial and final therapeutic complexity (Columns C*D) and look at the significant differences greater than 5%, we see that the final complexity increases in the physical sub-areas (54.6% vs 49.5%; p<0.001), practice (42.2% vs 37.1%; p<0.001)) and circumstances of the last days (16.5% vs 7.5%; p<0.001).

The analysis of the sub-areas of the HexCom of the lost patients showed some differences compared to the 1,555 with complete follow-up (Columns B*C): in general, their complexity was lower in some sub-areas of the psychic, spiritual, social and family, ethical and death-related areas (Table 3).

#### Multivariate analysis

Table 4 summarizes the bivariate Poisson regression analysis in order to explore the variables to be considered as risk factors of medium-high therapeutic complexity, at the initial and final moments. Those that showed a significant association were considered as variables for inclusion.

**Table 4.** Bivariate Logistic Regression Analysis with items from the Initial HexCom (A) and the Final HexCom (B)

| Variable | Initial HexCom Therapeutic Complexity (A) |  |  | Final HexCom Therapeutic Complexity (B) |  |  |
| --- | --- | --- | --- | --- | --- | --- |
|  | Beta coef | OR (CI95%) | p | Beta coef | OR (CI95%) | p |
| **<u>Socio-demographic/clinical</u>** |  |  |  |  |  |  |
| Age >80 years old | -0.219 | 0.804 (0.65 - 0.99) | 0.039 | -0.407 | 0.666 (0.54 - 0.82) | <0.001 |
| Sex | 0.092 | 1.096 (0.89 - 1.35) | 0.387 | 0.080 | 1.084 (0.88 - 1.34) | 0.453 |
| Caregiver partner | 0.113 | 1.120 (0.91 - 1.38) | 0.286 | 0.095 | 1.100 (0.89 - 1.36) | 0.373 |
| External caregiver | -0.096 | 0.909 (0.72 - 1.14) | 0.411 | -0.164 | 0.848 (0.67 - 1.07) | 0.165 |
| Death at home | -0.141 | 0.868 (0.70 - 1.08) | 0.203 | -0.639 | 0.528 (0.43 - 0.66) | <0.001 |
| Impairment (Pfeiffer) | -0.147 | 0.863 (0.69 - 1.08) | 0.205 | -0.221 | 0.802 (0.64 - 1.01) | 0.061 |
| Barthel |  |  |  |  |  |  |
| Independent patient | reference |  |  | Reference |  |  |
| Slight dependence | 0.207 | 1.229 (0.67 - 2.27) | 0.509 | 0.462 | 1.588 (0.89 - 2.84) | 0.119 |
| Moderate dependence | 0.331 | 1.393 (1.00 - 1.95) | 0.052 | 0.060 | 1.062 (0.77 - 1.46) | 0.715 |
| Severe dependence | 0.249 | 1.282 (0.92 - 1.80) | 0.149 | -0.062 | 0.940 (0.68 - 1.30) | 0.708 |
| Total dependence | -0.032 | 0.968 (0.68 - 1.39) | 0.86 | -0.568 | 0.567 (0.40 - 0.81) | 0.002 |
| Disease group |  |  |  |  |  |  |
| Oncological disease | reference |  |  | Reference |  |  |
| Advanced chronic organ failure | 0.177 | 1.193 (0.89 - 1.59) | 0.232 | 0.069 | 1.071 (0.79 - 1.46) | 0.662 |
| Advanced chronic neurological disease | 0.144 | 1.155 (0.71 - 1.88) | 0.565 | 0.533 | 1.705 (1.04 - 2.79) | 0.033 |
| Dementia | -0.214 | 0.807 (0.54 - 1.21) | 0.296 | -0.464 | 0.629 (0.41 - 0.96) | 0.031 |
| Frailty/geriatric multi-morbidity | 0.192 | 1.211 (0.67 - 2.18) | 0.523 | -0.196 | 0.822 (0.44 - 1.53) | 0.537 |
| **HexCom items** |  |  |  |  |  |  |
| Physical | 1.761 | 5.820 (4.60 - 7.37) | <0.001 | 2.039 | 7.680 (5.95 - 9.92) | <0.001 |
| Emotional | 0.925 | 2.521 (2.04 - 3.11) | <0.001 | 1.228 | 3.415 (2.75 - 4.25) | <0.001 |
| Personality | 0.961 | 2.614 (2.08 - 3.29) | <0.001 | 1.015 | 2.759 (2.18 - 3.50) | <0.001 |
| Transcendence | 1.038 | 2.823 (2.19 - 3.63) | <0.001 | 1.160 | 3.190 (2.47 - 4.11) | <0.001 |
| Connection | 1.378 | 3.965 (2.84 - 5.54) | <0.001 | 1.390 | 4.015 (2.85 - 5.66) | <0.001 |
| Meaning | 0.847 | 2.334 (1.84 - 2.95) | <0.001 | 0.939 | 2.556 (2.01 - 3.25) | <0.001 |
| Relational | 1.317 | 3.733 (2.93 - 4.75) | <0.001 | 1.618 | 5.045 (3.90 - 6.52) | <0.001 |
| Social and family emotional | 1.004 | 2.729 (2.21 - 3.37) | <0.001 | 1.248 | 3.483 (2.80 - 4.33) | <0.001 |
| Practice | 1.233 | 3.430 (2.77 - 4.25) | <0.001 | 1.397 | 4.041 (3.24 - 5.03) | <0.001 |
| External | 0.348 | 1.416 (1.15 - 1.74) | 0.001 | 0.309 | 1.362 (1.11 - 1.68) | 0.004 |
| Economic | 0.582 | 1.790 (1.36 - 2.36) | <0.001 | 0.528 | 1.695 (1.28 - 2.24) | <0.001 |
| Desire to die | 0.798 | 2.221 (1.51 - 3.27) | <0.001 | 0.724 | 2.063 (1.40 - 3.04) | <0.001 |
| Clinical decisions | 0.872 | 2.392 (1.85 - 3.09) | <0.001 | 1.038 | 2.824 (2.16 - 3.69) | <0.001 |
| Information | 1.057 | 2.877 (2.15 - 3.85) | <0.001 | 1.635 | 5.130 (3.56 - 7.40) | <0.001 |
| Location | 0.493 | 1.638 (1.33 - 2.02) | <0.001 | 0.960 | 2.613 (2.10 - 3.25) | <0.001 |
| Circumstances of the last days | 1.735 | 5.671 (3.79 - 8.48) | <0.001 | 1.229 | 3.418 (2.59 - 4.51) | <0.001 |
| Bereavement | 0.685 | 1.983 (1.60 - 2.46) | <0.001 | 0.682 | 1.978 (1.60 - 2.45) | <0.001 |

Two resulting multivariate models were obtained, one for initial and one for final therapeutic complexity (Table 5).

**Table 5.** Results of the multivariate analysis of the sub-areas of the HexCom determinants of therapeutic complexity at the end of life.

| 6 Main areas of HexCom* | Initial HexCom Evaluation |  |  |  | Final HexCom Evaluation |  |  |  |
| --- | --- | --- | --- | --- | --- | --- | --- | --- |
|  | Related Sub-areas* | Beta coefficient | OR (CI95%) | p | Related Sub-areas* | Beta coefficient | OR (CI95%) | p |
| **Clinical** |  |  |  |  |  |  |  |  |
|  | *Physical* | 1.517 | 4.56<br>(3.55 ; 5.86) | <0.001 | *Physical* | 1.775 | 5.90<br>(4.47 ; 7.79) | <0.001 |
| **Social and Family** |  |  |  |  |  |  |  |  |
|  | *Relational* | 0.599 | 1.82<br>(1.35 ; 2.45) | <0.001 | *Relational* | 1.046 | 2.85<br>(2.06 ; 3.92) | <0.001 |
|  | *Practice* | 0.624 | 1.87<br>(1.45 ; 2.41) | <0.001 | *Practice* | 0.852 | 2.34<br>(1.75 ; 3.15) | <0.001 |
|  |  |  |  |  | *External* | -0.516 | 0.60<br>(0.45 ; 0.80) | 0.001 |
| **Death** |  |  |  |  |  |  |  |  |
|  | *Circumstances of the last days* | 0.962 | 2.62<br>(1.62 ; 4.22) | <0.001 | *Circumstances of the last days* | 0.699 | 2.01<br>(1.43 ; 2.82) | <0.001 |
| **Ethics** |  |  |  |  |  |  |  |  |
|  |  |  |  |  | *Clinical decisions* | 0.454 | 1.58<br>(1.13 ; 2.21) | 0.008 |
|  | *Information* | 0.468 | 1.60<br>(1.12 ; 2.28) | 0.010 | *Information* | 0.711 | 2.04<br>(1.29 ; 3.23) | 0.002 |
| **Psycho-emotional** |  |  |  |  |  |  |  |  |
|  | *Personality* | 0.618 | 1.86<br>(1.41 ; 2.44) | <0.001 | *Personality* | 0.605 | 1.83<br>(1.36 ; 2.46) | <0.001 |
| **Spiritual** |  |  |  |  |  |  |  |  |
|  | *Meaning* | 0.289 | 1.34<br>(1.01 ; 1.76) | 0.041 |  |  |  |  |
|  | Constant | -2.442 |  |  | Constant | -2.502 |  |  |

\* See Table 1

The initial therapeutic complexity was more strongly related to the existence of difficulties related to the clinical area of HexCom (“physical” sub-area) (physical discomfort due to symptoms and/or injuries) (OR 4.56) and also to the death-related area of HexCom (sub-area “circumstances of the last days”) (difficulties in handling the dying process), with (OR 2.61). Then, with the social and family area of HexCom (“practice” sub-area) (difficulties with basic needs such as hygiene and food) (OR 1.86), with the psycho-emotional area (“personality” sub-area) (psychological vulnerability with difficulties adapting to changes) (OR 1.85) and with the social and family area (“relational” sub-area) (difficulties in relationships within the family environment) (OR 1.82). Finally, the initial therapeutic complexity was also significantly related to problems related to information (diagnosis and/or prognosis) (ethical area of HexCom, sub-area “information”) and to the existence of discomfort in the spiritual area (spiritual area of HexCom, sub-area “meaning”) (OR 1.59 and 1.33, respectively).

On the other hand, with regard to factors associated with the final therapeutic complexity, unlike the determining factors in the initial model, in this case we find that the spiritual complexity in the meaning sub-area ceases to be significant and the discomfort due to the lack of effective external support in the nucleus that lives together (Social and Family dimension, “external” sub-area) (OR 0.60) and difficulties in making clinical decisions (adequacy of diagnostic and/or therapeutic effort) (Ethical dimension, “clinical decisions” sub-area) (OR 1.58) becomes significant. In Social and Family complexity, “external” sub-area, for each unit that increases discomfort due to the lack of effective support external to the nucleus that lives together, the therapeutic complexity decreases, while the presence of the other variables increases the therapeutic complexity.

## Discussion

As main results of the present study, three facts should be highlighted: 1 - Some 31.6% of patients cared for by the *PADES* teams showed therapeutic complexity at the beginning of the process and this proportion increased to 35.5% at the end. In this regard, it can be said that a little more than a third of patients at this end-of-life stage may have therapeutic complexity at some point during the process. 2 - This therapeutic complexity both at the beginning and at the end is strongly related to clinical aspects (sub-area “physical”) such as the management of symptoms and to aspects related to the process of dying (sub-area “circumstances of the last days”). 3 - The therapeutic complexity increases as the process approaches the last days of life and death. This is probably due to a greater need for support and care, at a time when emotional stress due to the proximity of loss is higher. This could make relatives and caregivers perceive these moments as the most difficult in a global way. This would explain the fact that therapeutic complexity is related to almost all areas of complexity evaluated in the HexCom instrument, as shown in Table 4 of this study. Finally, it should also be noted that therapeutic complexity was significantly related to patients who were younger (<80 years old), more independent (better values in the Barthel index) and with advanced neurological disease (dementia excluded). This could be due to the fact that being younger and more independent can lead to a greater need for support and a greater emotional impact on the family environment.

Social and family complexity (relational, practical and external sub-areas). In this way, several studies have highlighted different aspects related to the needs of carers for the correct administration of medicines in end-of-life situations [21–22], as well as in pain management [23–24]. Therapeutic complexity does not only include access to drugs or techniques, but also adherence [25] to pharmacological treatment, which could translate into the acceptance of this treatment or the disparity between what the patient [26], relative or carer expects from this treatment and what will actually be achieved by following the recommendations, where sometimes, this reluctance comes from the lack of effective information on the part of the professionals who care for the patient [22–23]. At this moment, the technical aspect is not relevant if the social and family, spiritual or ethical factors are.

We must bear in mind that the circumstances at the end of life, and as we can see in our cohort, where more than 70% of patients with therapeutic complexity have some degree of dependence and 30% cognitive impairment, relatives and carers are often the ones who communicate with the doctor and express needs, so a disagreement with the treatment on the part of the relative can lead to low adherence to the treatment, which leads to poor control of symptoms and therefore an increase in clinical complexity. An example of this fact can be found in two published studies [23–24], which highlight the low adherence to opioid treatment due to mistrust or doubts about opioid treatment on the part of family members or caregivers, a fact that caused not only poor pain control but also a barrier to the use of analgesics by the patient. Poor pain control due to low adherence to pharmacological treatment limits recommendations or interventions by healthcare professionals to respond to the health problem that is being considered, which means that this problem, possibly “simple” to solve, becomes complex. Once translated into complexity, the tools available to resolve the patient’s clinical complexity are limited, a factor strongly related to therapeutic complexity.

Another important aspect is the role of the carer in this whole process, who not only intervenes in communication with the healthcare team that supports the patient but who also has to manage aspects such as the administration of medication (a complex issue for them) and be able to manage medication, patient care needs, management of visits, all added to the family role that this family member or carer may already have in their family, work and social environment, where the fact of being in charge of patient care can be burdensome [27–29], in addition to the significant therapeutic burden [2,22,23,30]. The descriptive study published by van Ryn et al. [31] based on interviews with patients and informal carers (mostly family members of the patient), roughly quantifies in half a working day what it means to take care of a patient with terminal stage cancer. At a time of high therapeutic burden, the family member/caregiver can be overwhelmed by the situation, a fact that can be reflected in the decisions that must be made (treatments, administration of medication, etc.) [32–31]. This fact agrees with the results obtained in our study, where the therapeutic complexity at the beginning of the process shows that difficulty in handling information about the diagnosis and the prognosis increases therapeutic complexity and in the final assessment of this complexity, difficulty making clinical decisions related to the adequacy of the diagnostic and/or therapeutic effort is added as a determining factor of complexity. The fact that in the initial assessment we only find difficulty in handling the information and in the final assessment we add difficulty in the adequacy of the therapeutic effort, we believe is caused by the process of the evolution of the disease itself towards the circumstances of the last days. Thus, at the beginning of the process, the impact of the information on the prognosis is much more relevant than when it reaches the last days, where the making of consensual decisions between the patient, the family and the team becomes more difficult and is driven by time pressure (time running out).

The management of medication at the end of life is management of a complex regime [5,33], due to overload and the fact of having to take charge of relevant aspects in patient care which are related to prognosis such as: 1) decision-making with regard to adequacy of therapeutic effort, especially in the phase of the last days, 2) administration of medications to avoid hospital admission, 3) storage of medications at home, 4) fear of administration of an accidental overdose, 5) fear that administration of a drug will advance the death process, 6) understanding aspects related to poly-medication and, finally, 7) when to use one drug or the other according to symptomatology. All this is added to the fact that often the taking or administration of different medicines does not match the schedule or route of administration, making it even more difficult to manage the medication and therefore therapeutic complexity [21].

The study published by Wilson et al. [29], identifies that, with regard to the work associated with the management of medication in the patient at the end of life at home, practical, physical, emotional and knowledge elements are included that involve both caregivers/family members and patients to manage prescriptions for multiple symptoms, different and often highly variable administrations in a short period of time and which therefore adds a considerable burden when someone is cared for at home in an end-of-life process. The results of our study would agree with those where the therapeutic complexity pivots, to a greater or lesser extent, around the six areas described in HexCom: clinical, spiritual, social and family, ethical and those related to death or the process of dying.

Complexity in patients undergoing palliative care is a fact that many authors highlight and is collected in multiple articles, where different instruments have also been developed in order to be able to understand it [12]. Despite the lack of a formal definition of what complexity is at the end of life [34–35], all articles refer to this complexity as the result of the interrelationship of the patient’s needs through multiple dimensions of needs (psychological, physical and spiritual needs, among others), where this complexity lies in the non-linearity of these interactions [36] and to which the dynamic aspect of the constant change in the patient’s state is added.

## Strengths and limitations

Possible limitations of the study may include bias since in the initial design of the study the factors or combination of factors were not collected, which marked the consensus of the professionals as moderate-high, moderate or low complexity. It should also be considered that variables specific to the treatment were not included (number of medications, active principle, route of administration, etc.) that could also influence the therapeutic complexity. Although we consider that in future studies these aspects should be included, in an indirect way this fact was taken into account in the assessment of the therapeutic complexity that was made by consensus among the *PADES* team that supported the patient. Another aspect to consider are the losses in the final evaluation of the patients included in the study cohort that may affect the results of the analysis of the factors that influence the final therapeutic complexity. Although the percentage of losses is low, we do see in the analysis that there are differences between the patients who ultimately remain in the cohort and those who did not manage to carry out this final assessment. However, these lost patients, in general, show less complexity in some sub-areas of HexCom, so we can assume that the final analysis is representative of the studied cohort.

As a strength of the study, we must consider that it is a large cohort of patients with a large sample. Also the fact that it is a multi-centre prospective study, with longitudinal data. It should also be noted that the assessment of therapeutic complexity is carried out by consensus of an interdisciplinary team, always including medical, nursing and social work professionals and the fact of having carried out previous training limits the systematic bias in the collection of information. Considering the results, we can say that there is a concordance of the results obtained with the literature. All the factors that determine therapeutic complexity evaluated using the HexCom model are in line with the results of the different studies consulted [12,29,34–36]. Finally, it should also be mentioned that the results obtained are consistent, from a clinical point of view and agree with the experience of what is considered normal clinical practice.

## Conclusion

In palliative home care, therapeutic complexity is related to difficulty in managing symptoms or the circumstances of the last days and, in parallel, it is related to difficulty in the suitability of the therapeutic effort. It is also related to social and family complexity (difficulties in family relationships, handling the patient’s basic needs, and the lack of external support for the family nucleus).

This implies that not only giving valid instructions for the correct use of medication will achieve effective management, but it is also necessary to provide support in other areas, such as social and family and ethical areas, in order to reduce therapeutic complexity for patients with palliative care needs.

## Data Availability

Since this study is a national study, we did not apply for permission to share the datasets with external researchers outside the country. Hence, this was not included in the consent forms that were signed by patients. Some data may be available upon request.

## Acknowledgements

We would like to express our gratitude to the study nurses, and clinicians who participated as well as to all patients who were included.

The authors thank Metzger translations for English language support.

